# Artificial intelligence-generated smart impression from 9.8-million radiology reports as training datasets from multiple sites and imaging modalities

**DOI:** 10.1101/2024.03.07.24303787

**Authors:** Parisa Kaviani, Mannudeep K. Kalra, Subba R Digumarthy, Karen Rodriguez, Sheela Agarwal, Rupert Brooks, Sovann En, Tarik Alkasab, Bernardo C. Bizzo, Keith J. Dreyer

**Affiliations:** Department of Radiology, Massachusetts General Hospital and Harvard Medical School, Boston, MA; MGH & BWH Center for Clinical Data Science, Boston, MA, USA; Nuance Communications, Burlington, MA, USA; Nuance Communications, Montreal, QC, Canada; CADT (Cambodia Academy of Digital Technology), Bridge 2, National Road 6A, Sangkat Prek Leap, Khan Chroy Changva, Phnom Penh

## Abstract

**Importance:** Automatic generation of the impression section of radiology report can help make radiologists efficient and avoid reporting errors.

**Objective:** To evaluate the relationship, content, and accuracy of an Powerscribe Smart Impression (PSI) against the radiologists’ reported findings and impression (RDF).

**Design, Setting, and Participants:** The institutional review board approved retrospective study developed and trained an PSI algorithm (Nuance Communications, Inc.) with 9.8 million radiology reports from multiple sites to generate PSI based on information including the protocol name and the radiologists-dictated findings section of radiology reports. Three radiologists assessed 3879 radiology reports of multiple imaging modalities from 8 US imaging sites. For each report, we assessed if PSI can accurately reproduce the RDF in terms of the number of clinically significant findings and radiologists’ style of reporting while avoiding potential mismatch (with the findings section in terms of size, location, or laterality). Separately we recorded the word count for PSI and RDF. Data were analyzed with Pearson correlation and paired t-tests.

**Main Outcomes and Measures:** The data were ground truthed by three radiologists. Each radiologists recorded the frequency of the incidental/significant findings, any inconsistency between the RDF and PSI as well as the stylistic evaluation overall evaluation of PSI. Area under the curve (AUC), correlation coefficient, and the percentages were calculated.

**Results:** PSI reports were deemed either perfect (91.9%) or acceptable (7.68%) for stylistic concurrence with RDF. Both PSI (mismatched Haller’s Index) and RDF (mismatched nodule size) had one mismatch each. There was no difference between the word counts of PSI (mean 33±23 words/impression) and RDF (mean 35±24 words/impression) (p>0.1). Overall, there was an excellent correlation (r= 0.85) between PSI and RDF for the evolution of findings (negative vs. stable vs. new or increasing vs. resolved or decreasing findings). The PSI outputs (2%) requiring major changes pertained to reports with multiple impression items.

**Conclusion and Relevance:** In clinical settings of radiology exam interpretation, the Powerscribe Smart Impression assessed in our study can save interpretation time; a comprehensive findings section results in the best PSI output.

## Introduction

The workforce is the pillar of any industry and impacts how products and services reach consumers. For the healthcare industry, in particular, workflow affects the timely delivery of critical services that can make a big difference in patients’ lives. The ongoing COVID-19 pandemic has brought workforce challenges to the center stage, cutting across most industries including healthcare. Not unlike other medical specialties, radiology also suffers from workforce shortage issues related to both technologists and radiologists [1]. As hospitals and departments search for additional hires, the current workforce deals with the pressure to provide more services to meet the demands of a population suffering either from COVID-19 or other ailments that took a backseat during the pandemic [2]. According to a recent survey of 254 hospitals from the United States (US), 45% of hospitals had staff shortages in their radiology departments contributing to inadequate healthcare services [3]. The Association of American Medical Colleges estimates a shortage of 42,000 radiologists and clinicians by the year 2033.

Advances in imaging technologies in the past two decades have contributed to a tremendous increase in imaging utilization. The increasing utilization of imaging further compounds the ongoing workforce shortage. Smith-Bindman et al reported a substantial surge in CT, MRI, and ultrasonography from 56, 16, and 177 per 1000 individuals in the year 2000 to 141, 64, and 347 per 1000 individuals in 2016, respectively [4]. In addition to the burgeoning imaging volume, technological capabilities have also increased the volume of data and images generated from individual imaging examinations. Routine juxta- or sub-millimeter section thickness with multiplanar acquisitions or reformats are now common and recommended for several clinical indications such as lung nodule follow-up, lung cancer screening, CT angiography, and diffuse lung diseases.

Interpreting physicians review images and create radiology reports often structured in two to four sections: imaging procedure or protocol name, findings, impression, and recommendation sections. The impression section abstracts the salient findings and presents possible diagnosis or differential diagnosis, as well as disease evolution from prior imaging when available. Apart from the additional dictation time, the impression section can also contain errors or discrepancies from the details in the findings section such as those related to omission (key details in the finding section not summarized or addressed in the impression) or commission (errors of laterality, disease evolution, or distribution). Therefore, we hypothesized that radiology report impressions generated with a fully automatic, artificial intelligence (AI) algorithm could help save interpretation time and avoid errors and discrepancies. The purpose of our study was to compare the Powerscribe Smart Impression (PSI) section of radiology reports and radiologists-dictated impression (RDF).

## Material and Methods

### Ethical approval and disclosures

Our retrospective study was approved by the institutional review board at Massachusetts General Brigham (IRB protocol number: 2020P003950) with a waiver of informed consent. The study followed the Health Insurance Portability and Accountability Act’s guidelines (HIPAA). Three coauthors (SA, RB, and SE) are employees of Nuance Communications. Two study coinvestigators (MKK and SRD) have received research grant funding for unrelated projects (Coreline Inc., Riverain Tech, Siemens Healthineers; Qure.AI, Lunit Inc., Vuno Inc.). There was no research grant, fund, or support provided for this study.

### Exam types and Radiologists

The study included 3879 radiology reports of imaging exams performed in adult patients (>18 years). These reports included 2798 CT, 502 radiographs, 345 ultrasound exams, 116 MRI, 88 PET CT and 30 other imaging tests, all performed between February 2022 and April 2022. Of these 3879 reports, 2114 reports were randomly selected from eight radiologists (five thoracic and three abdominal radiologists) at a quaternary care hospital (Massachusetts General Hospital). From this site, 1358 reports from the five thoracic radiologists included 833 chest CT, 1 MRI, 479 chest radiographs, 40 PET-CT, 3 ultrasonography, and 2 fluoroscopic procedures. The 756 radiology reports from the three abdominal radiologists included 240 abdomen-pelvis CT, 75 MRI, 23 radiographs, 342 ultrasonography, 48 PET-CT, and 28 other imaging tests (such as upper gastrointestinal series and other fluoroscopic procedures).

The remaining 1765 reports (1140 abdominal radiology reports and 625 thoracic radiology reports) were randomly selected from 8 imaging sites representing a mix of hospitals and outpatient imaging centers across the United States.

Three radiologists (SRD, MKK, KR with 18, 15, and 3 years of experience as interpreting radiologists) reviewed the radiology reports to record the number of incidental and significant findings in the RDF and PSI. In addition, they assessed both the RDF and PSI for stylistic concordance and any discordance with the findings section. Upon review, the radiologists excluded two radiology reports without any finding and impression sections. Thus, the final sample size was 3879 radiology reports.

### AI Model and data partitioning

We used a neural abstractive summarization approach to generate the impressions in this research study. All report sections other than the impression, supplemented with certain basic metadata, were used as the input. A collection of radiology reports from ten different healthcare centers in the United States was used as training data. Duplicate, or very near duplicate reports, as well as any radiology reports without an impression section were excluded from training. The data were prepared by normalizing and word tokenizing the radiology report text. We also excluded reports with more than 512 tokens or which had an impression section with more than 128 tokens. This process led to a set of 9.8 million usable reports for training. As part of the tokenization process, all dates and times were reduced to generic “_date_” and “_time_” tags. We used a sequence-to-sequence transformer [5] with pre-layer normalization [6]. The system is trained from scratch using teacher forcing and cross-entropy loss. The vocabulary size was 33,000 and the encoder and decoder have 6 layers resulting in 64 million parameters for the entire model.

### Evaluation

A separate set of radiology reports was imported into a Microsoft Access file (Microsoft Office 2016) to create a user-friendly evaluation and data entry system in which the data were presented in a desired structure (Figure 1). The imported dataset in the Microsoft Access interface displayed the original finding section, RDF, and PSI, as well as word counts for both impressions. In addition, the interface allowed radiologists to record the following parameters: separate numbers of incidental and significant findings in the RDF and PSI; finding progression (new or increasing, stable, decreased or resolved findings) in both RDF and PSI; any lack of consistency between the finding section and the RDF and PSI; stylistic concordance of PSI with the RDF (excellent, acceptable, suboptimal); and the overall evaluation of PSI on a four-point scale (1: excellent without need for any changes; 2: Good with minor changes or tweaks; 3: some major changes needed, but the PSI was still helpful; 4: unacceptable, with too many changes or mistakes). The interface also provided a free text box to enter specific comments on any aspect of PSI or RDF. Figure 1 is a snippet of Microsoft Access for reviewing an example radiology report.

**Figure 1.**
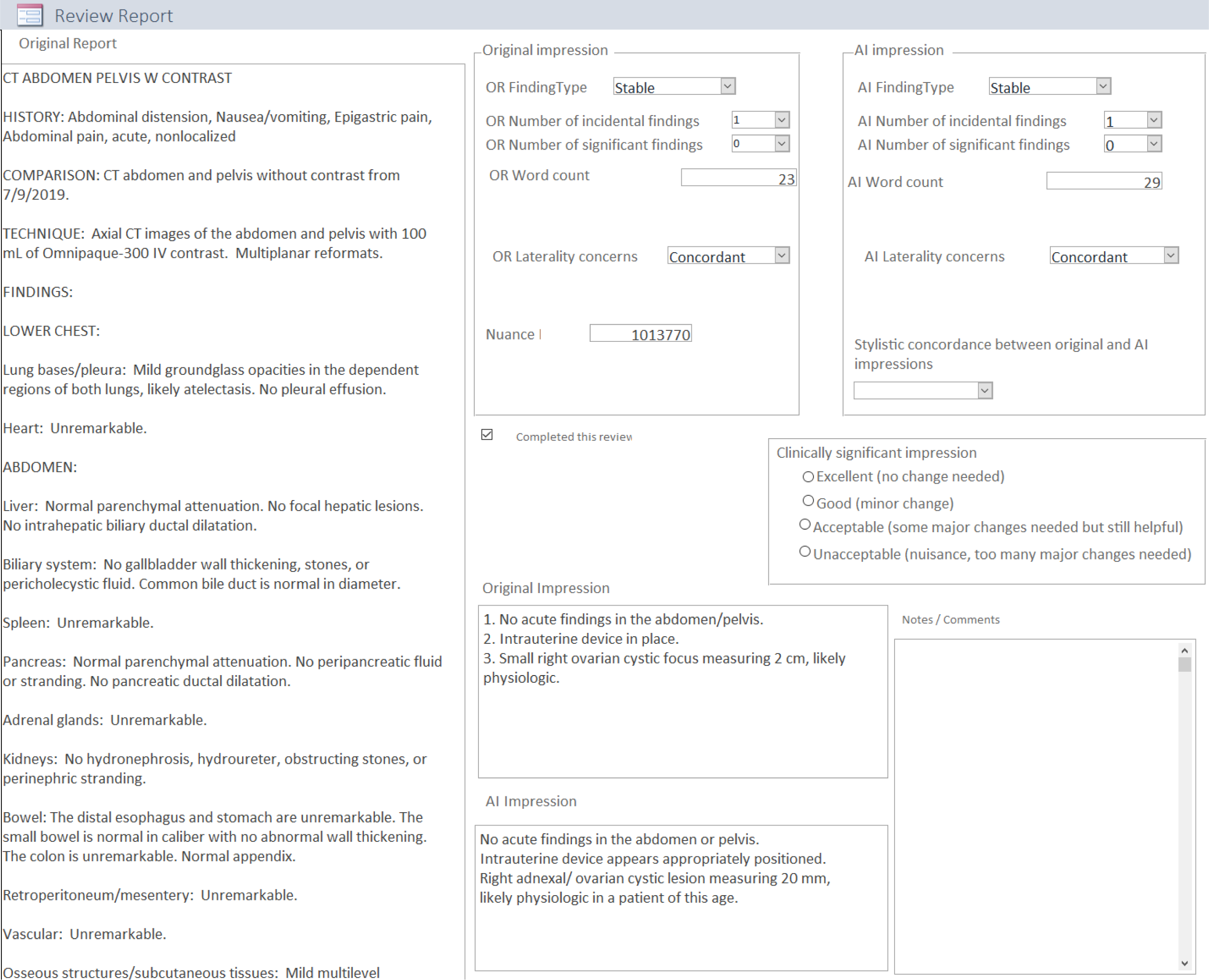
Screenshot of the Microsoft Access interface created for reviewing radiology reports.

### Statistical Evaluation

We performed statistical analysis with Microsoft EXCEL 16 (Microsoft Inc., Redmond, Washington) and SPSS version 26 (IBM Inc., Chicago, Illinois). The frequency of the incidental and significant findings was recorded in the range of zero to nine. If the number of the findings was equal to or greater than ten, it was recorded as ≥10. To assess the AI performance, we defined true positive (RDF and PSI with an equal number of incidental and significant findings [total number of findings> zero]), true negative (no reported incidental and significant findings), false positive (PSI with one or more findings that were absent in the findings section and the RDF), and false negative (PSI without one or more findings documented in the RDF and findings section). We calculated sensitivity, specificity, and the area under the curve (AUC) for the receiver operating characteristics (ROC) analysis. Separately, we estimated the linear correlation coefficient between the word counts for RDF and PSI.

## Results

There were no significant differences between the word counts of RDF and PSI in both the single center reports (RDF: 33.0±24 words; PSI 30.7±23) and the multicenter reports (RDF: 38±24 words; PSI 36±23) (p>0.1). Likewise, both RDF and PSI were consistent with the original findings section of reports from both the single center [RDF: 98.1% (2074/2114), PSI: 98.8% (2085/2114)] and the multicenter data [RDF 98% (1736/1765), PSI: 99% (1749/1765)]. The frequency of discrepancies was significantly higher for RDF (single center: 40 reports; multicenter reports: 28) than for PSI (single center: 29 reports; multicenter reports: 16) (*p* <0.001). Table 1 summarizes the frequency of different types of discrepancies in both datasets.

**Table 1.**
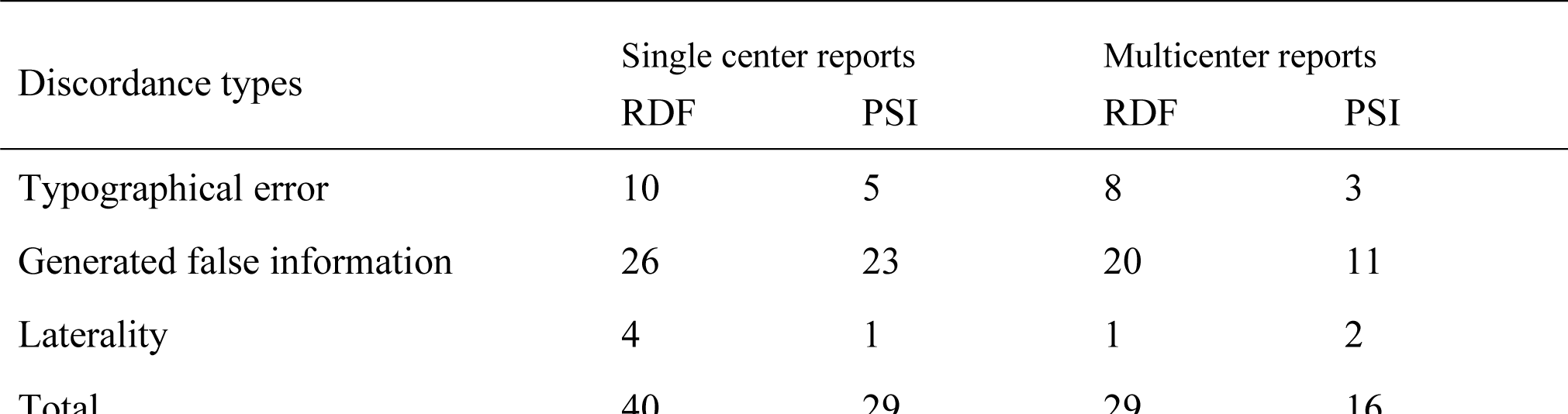
Summary of different types of recorded discrepancies in the single center and multicenter reports. The two laterality errors with PSI were both related to statement regarding right or left renal cysts rather than bilateral cysts. The false information emanated from discordance between the stated clinical indication for the imaging test in the findings section and in the RDF or PSI. (key: RDF-radiologist dictated impression, PSI-powerscribe smart impression)

There was perfect stylistic concordance between PSI and RDF in 98.3% and 83.9% of single-center and multicenter reports, respectively. There was a significantly higher correlation between the PSI and RDF for clinically significant findings (single center*, r*=0.887; multicenter reports, *r*=0.854) than for incidental findings (single center*, r*=0.717; multicenter reports, *r*=763) (*p*<0.001). Table 2 summarizes the frequency of the incidental and significant findings. There were no significant variations in the PSI performance or correlation with the RDF across radiology reports of different imaging modalities or across the eight radiologists from single center report datasets (p>0.1).

**Table 2.**
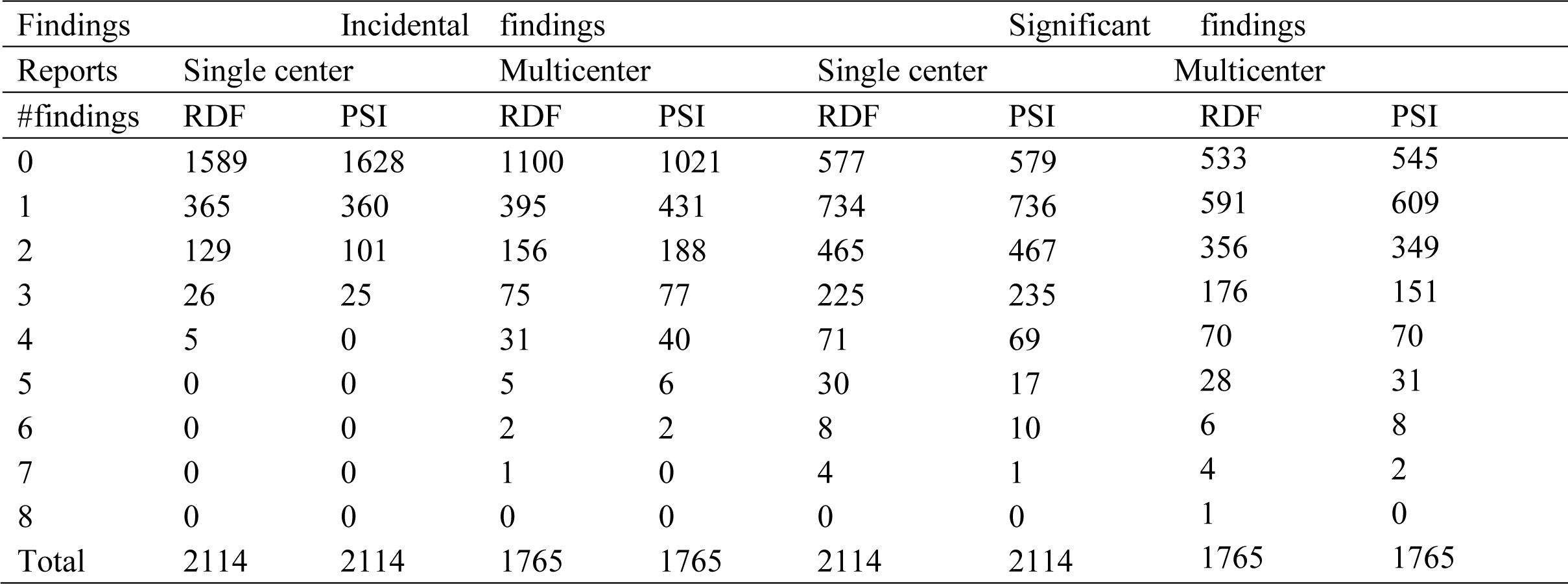
Summary of number of incidental and significant findings per report for both radiologist-dictated impression (RDF) and powerscribe smart impression (PSI).

| Findings | Incidental findings |  |  |  | Significant findings |  |  |  |
| --- | --- | --- | --- | --- | --- | --- | --- | --- |
| Reports | Single center |  | Multicenter |  | Single center |  | Multicenter |  |
| #findings | RDF | PSI | RDF | PSI | RDF | PSI | RDF | PSI |
| 0 | 1589 | 1628 | 1100 | 1021 | 577 | 579 | 533 | 545 |
| 1 | 365 | 360 | 395 | 431 | 734 | 736 | 591 | 609 |
| 2 | 129 | 101 | 156 | 188 | 465 | 467 | 356 | 349 |
| 3 | 26 | 25 | 75 | 77 | 225 | 235 | 176 | 151 |
| 4 | 5 | 0 | 31 | 40 | 71 | 69 | 70 | 70 |
| 5 | 0 | 0 | 5 | 6 | 30 | 17 | 28 | 31 |
| 6 | 0 | 0 | 2 | 2 | 8 | 10 | 6 | 8 |
| 7 | 0 | 0 | 1 | 0 | 4 | 1 | 4 | 2 |
| 8 | 0 | 0 | 0 | 0 | 0 | 0 | 1 | 0 |
| Total | 2114 | 2114 | 1765 | 1765 | 2114 | 2114 | 1765 | 1765 |

In the overall assessment versus the RDF, PSI was either perfect [single center: 66.3% (1401/2114), multicenter: 67.7% (1196/1765)] or good [single center: 24.1% (510/2114); multicenter reports 22.7% (402/1765)] for most radiology reports regardless of the imaging modality. In a minority of reports, PSI needed major changes [single center, 8.0% (169/2114); multicenter reports,6.8% (121/1765)] or was unacceptable [single center, 1.6% (34/2114); multicenter reports, 2.8% (46/1765)]. Table 3 summarizes the distribution of overall evaluation of PSI performance in terms of different modalities as well as different body parts. The AUCs for PSI were lower for incidental [single center reports: 0.75 (95% CI 0.722-0.779); multicenter 0.70 (95% CI 0.673-0.727)] than for clinically significant findings [single center, 0.76 (95% CI 0.732-0.787), multicenter 0.785 (95% CI 0.762-0.808)]. Figure 2 summarizes the distribution of discordance, stylistic concerns, and the overall evaluation of impression sections in the reports of different imaging modalities.

**Table 3.**
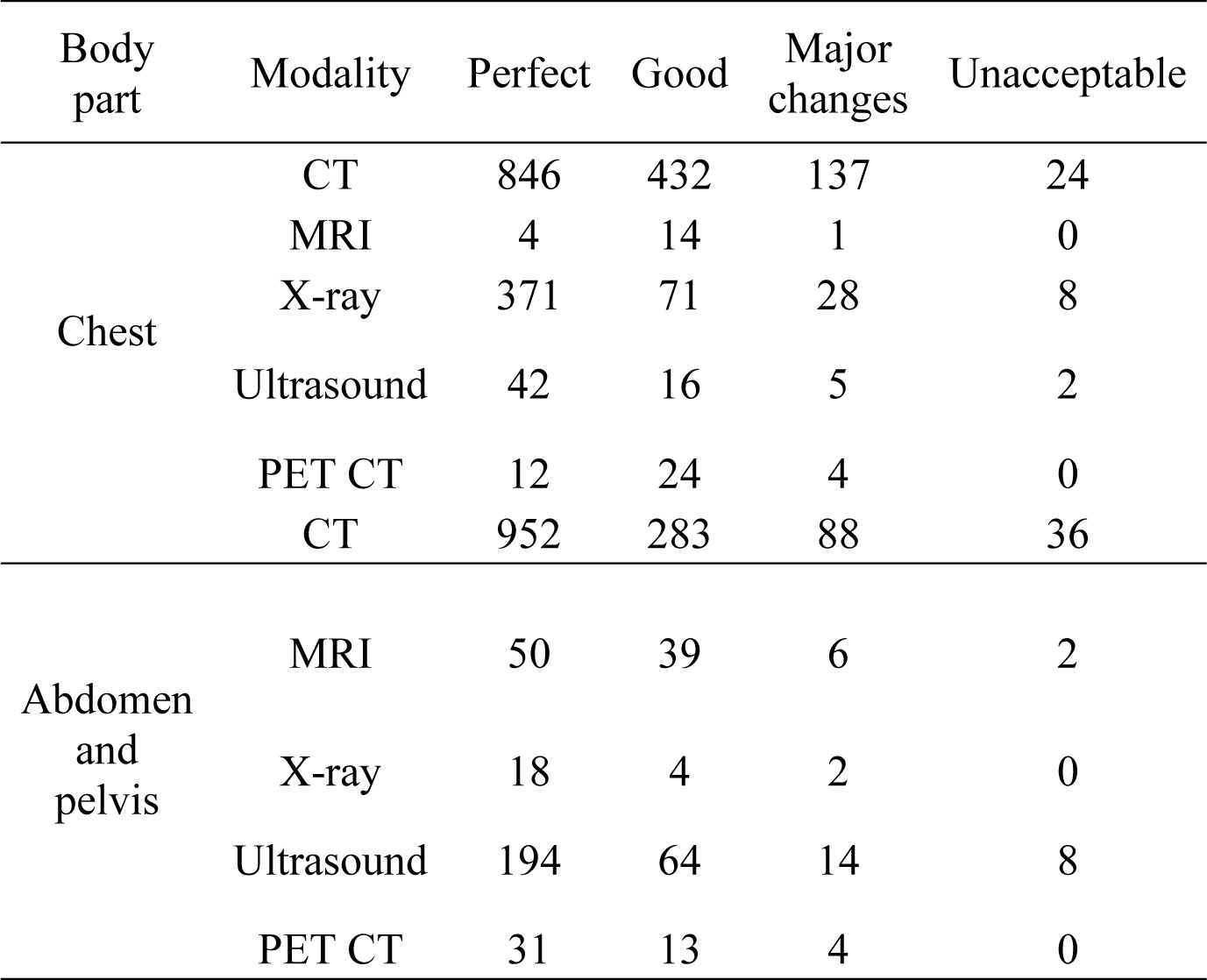
The overall evaluation of PSI in terms of the body parts as well as different modalities.

| Body part | Modality | Perfect | Good | Major changes | Unacceptable |
| --- | --- | --- | --- | --- | --- |
| Chest | CT | 846 | 432 | 137 | 24 |
|  | MRI | 4 | 14 | 1 | 0 |
|  | X-ray | 371 | 71 | 28 | 8 |
|  | Ultrasound | 42 | 16 | 5 | 2 |
|  | PET CT | 12 | 24 | 4 | 0 |
|  | CT | 952 | 283 | 88 | 36 |
| Abdomen and pelvis | MRI | 50 | 39 | 6 | 2 |
|  | X-ray | 18 | 4 | 2 | 0 |
|  | Ultrasound | 194 | 64 | 14 | 8 |
|  | PET CT | 31 | 13 | 4 | 0 |

**Figure 2.**
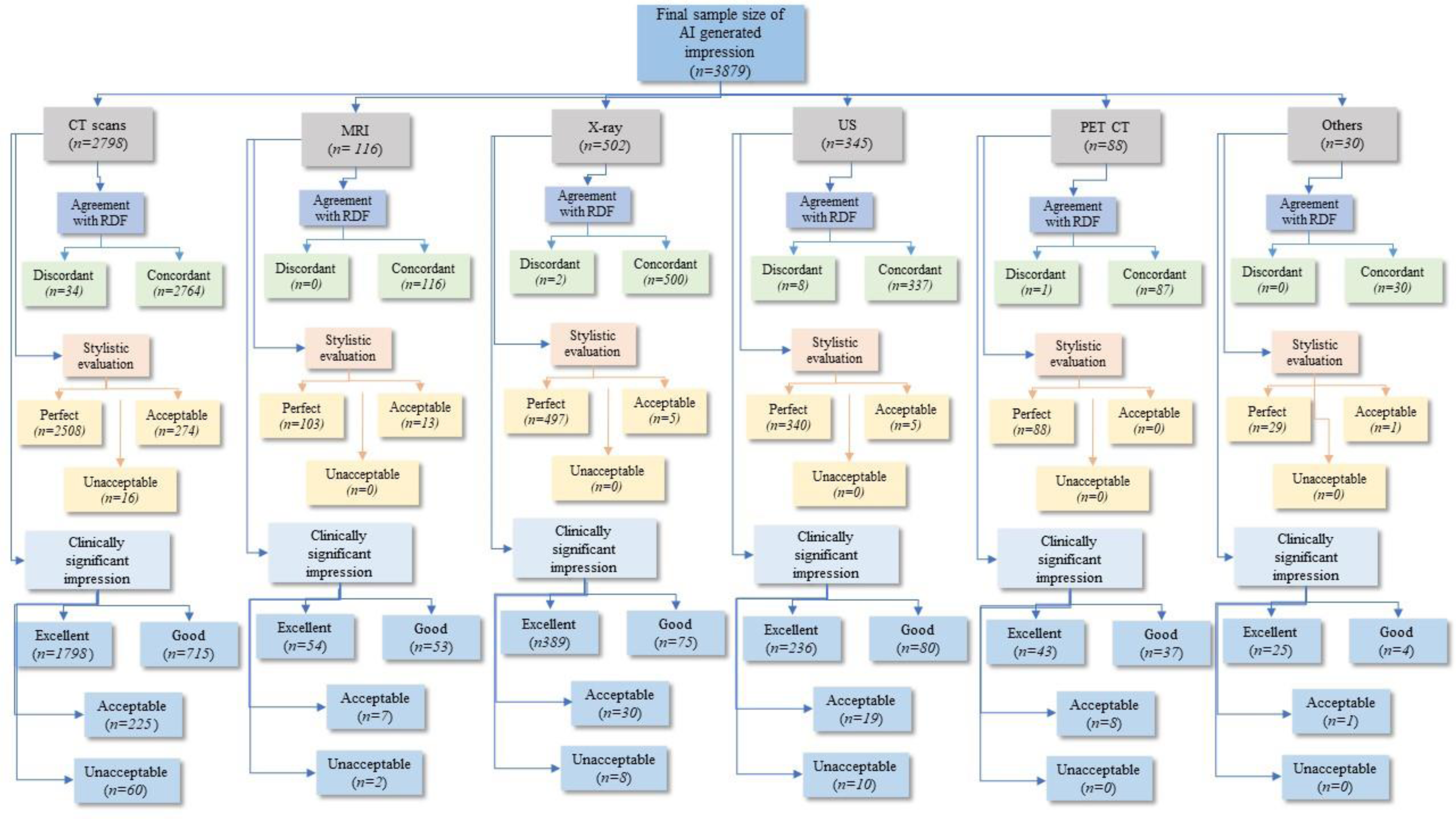
Summary of agreement with RDF, stylistic concerns, and clinically significant findings in the Powerscribe Smart Impression. (OR-original report)

## Discussion

We found high stylistic and content-based concordance between PSI and RDF. Our study highlights the potential of AI and natural language processing in automatically generating impression sections from the findings section.[7] Only a limited number of examples of generation of impression from the previous sections of radiology reports can be found in the literature. Zhang et. al. used an LSTM based encoder decoder approach [8]. Initial expert evaluation on 100 reports indicated that 67% of generated impressions were as good or better than the reference impression [9], but this figure was later improved to 72% by optimizing factual accuracy using reinforcement learning[10]. Sotudeh Gharebagh et al. [11] and Hu et al. [12] achieved more than 80% of generated impressions as good or better than human generated ones using more complex models. However, these results were limited to chest X-ray reports due to data availability. Macavaney et al. examined summarization of other report types, although limited to a single institution [13]. In each of these previous works, expert evaluation was done on a small set of 100 reports [9]. We are also aware of a separate clinical product that also generates an impression section from the physician-dictated findings section (https://www.radai.com accessed on December 1, 2022). Prior studies have used natural language processing and AI to create radiology reports text from images [7, 14, 15, 16].

PSI performed substantially better for findings deemed clinically significant than for incidental findings. This could be related to the inconsistent citation of incidental findings in the impression sections of radiologists or suggest a limitation of the PSI algorithm. It is likely that radiologists consistently summarize the significant findings in their impression sections but do not re-state incidental findings in the impression section to either keep the focus on significant findings or to be efficient with both verbiage and time. Another important aspect of our study is the lower rates of discordance between the findings section and PSI versus the finding section and RDF. Such RDF discrepancies have been reported in prior studies [17]. For example, Sangwaiya et al have evaluated the laterality mismatches and lesion side errors between the findings and impression sections [17]. Most errors (70.9%) in their study were deemed as clinically important. Lutemer et al have also reported on laterality errors in radiology reports both with and without the use of voice recognition for the interpretation of imaging exams [18]. We report fewer side discrepancies with PSI than with RDF.

Sangwaiya et al have also reported on the discrepancies in vertebral body levels of abnormalities described in the finding section with those summarized in the impression section [19]. Such errors can lead to the wrong surgery or intervention site with catastrophic impact. Although we did not specifically assess spinal MR or CT reports, we noticed no errors related to the numerical description of lesion size or vertebral level with the PSI.

The main implication of our study is the potential impact of PSI on reporting efficiency and discrepancies. The efficiency impact will be related to both the length of RDF and the accurate reflection of PSI. Shorter impressions such as “Clear lungs,” “Normal chest,” or “Negative study” are unlikely to benefit from the command “Macro powerscribe smart generator” during dictation or single-click on the shortcut menu in the dictation interface. In its clinical implementation, such a command or shortcut pops up a window containing PSI, which the radiologist can accept, reject, or edit as needed. Given the high accuracy of PSI, it is likely that PSI will be useful for longer, complex reports. Given the limitation of PSI in the absence of details about lesion evolution or differential diagnosis in the findings section, radiologists can modify their reporting patterns by including those aspects in the findings section. Although such report modification might improve PSI performance, it will likely decrease the overall efficiency benefits of PSI. On the other hand, the additional time to “strengthen” the findings section will improve the accuracy of PSI and avoid potential RDF errors (size, laterality, or missed mention) in complex reports. Since PSI does not operate in a default “always on” mode, radiologists can choose when to use the feature. Although PSI is trained with a large radiology report dataset from multiple sites, it personalizes the generated impression based on individual radiologists’ reports in the training data. Future studies will be needed to assess how PSI performs when structured impression sections are generated using clinical guidance tools such as for low-dose CT for lung cancer screening, mammography, and prostate MR.

There are several limitations to our study. First, the study represents an initial retrospective evaluation of PSI instead of real-world implementation and experience during formal, live interpretation of radiology examinations. However, due to the efforts and workflow implications, our AI governance body requires a formal investigation before any AI techniques are considered for clinical adoption in radiology workflow. Second, the study included imaging examinations from only two radiology subspecialties, and therefore, PSI’s performance across other body part examinations and reports cannot be commented upon. Third, there was an asymmetric distribution of imaging modalities among the assessed radiology reports with a preponderance of CT and radiography reports, and a smaller number of nuclear medicine, MR, ultrasound, and interventional procedures reports. Fourth, we did not intentionally enrich the evaluated reports to ensure a real-world scenario. There was an adequate representation of complex multi-finding reports in our study. Fifth, we also did not enrich our reports datasets with reports from radiologists with a “more verbose, literary, or descriptive style” of reporting. Sixth, although we evaluated reports belonging to 8 separate healthcare systems across the country, the adjudicating radiologists belonged to a single site. However, given the proclivity towards a more descriptive reporting style at the evaluating site is likely to yield PSI underperformance rather than an overperformance. Seventh, all evaluated reports were in the English language since the PSI is currently operational in English only. Eighth, we did not enrich specifically assess the PSI performance among imaging tests performed in inpatient, ICU settings, and outpatients. Nine, we did not assess PSI performance among reports or impressions dictated or completed using the clinical guidance tools for certain imaging protocols and findings (such as Lung Rads, TI-Rads, and LI-Rads). Such reports were represented in the evaluated datasets although their sample size did not reach an adequate number for drawing conclusions on PSI performance. Though none of the three evaluating radiologists are native English language speakers they had a combined radiology interpretation and reporting experience of more than 35 years in the United States. Finally, our study assessed the PSI in an offline, retrospective manner, and does not imply its clinical availability or performance in the real-world setting.

In conclusion, our study found that the PSI generates accurate impression section from radiology findings regardless of the reporting sites and imaging examination in thoracic and abdominal radiology subspecialties. As an aid tool, the AI generated impression can help improve reporting efficiency. Because the AI powerscribe smart impression generator trains itself on individual radiologists reporting patterns, the AI generated impression maintains a high level of stylistic concordance with the radiologists’ dictated reports. Furthermore, AI generated impressions can also help avoid common mistakes such as typographical errors, laterality discordance, and the frequency of clinically significant findings not mentioned in the impression section.

## Data Availability

N/A

